# The effects of alcohol use severity and polygenic risk on gray matter volumes in young adults

**DOI:** 10.1101/2025.01.20.25320842

**Authors:** Yu Chen, Huey-Ting Li, Xingguang Luo, Guangfei Li, Jaime S. Ide, Chiang-Shan R. Li

## Abstract

Genetic factors contribute to alcohol misuse. Chronic alcohol consumption is associated with decreases in gray matter volumes (GMVs) of the brain. However, it remains unclear whether or how genetic risks may alter GMVs independent of the effects of alcohol exposure. Here, we employed the Human Connectome Project data of neurotypical adults (n = 995; age 22-35; 618 women) and, with voxel-based morphometry analysis, computed the GMVs of 166 regions in the automated anatomical atlas 3. Alcohol use behaviors were assessed with the Semi-Structured Assessment for the Genetics of Alcoholism. Alcohol use severity was quantified by the first principal component (PC1) identified of principal component analysis of 15 drinking measures. Polygenic risk scores (PRS) for alcohol dependence were computed for all subjects using the Psychiatric Genomics Consortium study of alcohol dependence as the base sample. With age, sex, race, and total intracranial volume as covariates, we evaluated the relationships of regional GMVs with PC1 and PRS together in a linear regression. PC1 was negatively correlated with GMVs of right insula and Heschl’s gyrus, and PRS was positively correlated with GMVs of left posterior orbitofrontal cortex, bilateral intralaminar nuclei of the thalamus and lingual gyri. These findings suggest distinct volumetric neural markers of drinking severity and genetic risks of alcohol misuse. Notably, in contrast to volumetric reduction, the genetic risks of dependent drinking may involve larger regional volumes in the reward, emotion, and saliency circuits.

## 1. Introduction

Genetic factors influence individual susceptibility to alcohol misuse, accounting for more than 50% of the variance in alcohol use severity, as demonstrated in twin and adoption studies (Trudell et al., 2014, Ducci and Goldman, 2008, Verhulst et al., 2015). Polygenic risk scores (PRS), computed based on common genetic variants, predicted the severity of alcohol misuse (Savage et al., 2018, Taylor et al., 2016) and identified high-risk drinkers (Lai et al., 2022). However, the neural bases of the genetic risks of alcohol misuse remain largely unexplored. Characterizing the genetically informed neural phenotypes may help investigators distinguish the risks and consequences of misuse.

Chronic drinking is accompanied by widespread structural shrinkages of the frontal, parietal, temporal, and occipital lobes, the insula, as well as subcortical regions, including the amygdala, striatum, and thalamus (Yang et al., 2016, Zhornitsky et al., 2021, Luo et al., 2022b, Yang et al., 2024, Cao et al., 2021, Chye et al., 2020). The severity of drinking is correlated with lower regional gray matter volumes (GMVs) (Thayer et al., 2016, Mackey et al., 2019a) and functional loss in people with alcohol use disorder (AUD) (Li et al., 2021). For instance, lower medial frontal and insular GMVs were associated with worse cognitive and emotional functions in AUD (Spindler et al., 2021).

While this literature has described structural brain changes as the consequences of drinking, less research has aimed to identify brain markers of the genetic risks of alcohol misuse. An earlier study found larger bilateral insular surface area (SA) in AUD vs. healthy controls and identified 36 insular SA-related genes in significant correlation with AUD status with small effect sizes (*r*^2^ < 0.000001) (Zhao et al., 2019). Another study reported that alcohol-dependent individuals with two protective alleles of a single nucleotide polymorphism rs1789891 had reduced GMVs in bilateral superior, middle, and inferior temporal gyri (Bach et al., 2019). Luo and colleagues demonstrated significant and replicable association amongst KTN1 variants, KTN1 mRNA expression, and putamen GMVs in alcohol and comorbid substance misuse (Luo et al., 2021).

Given that AUD is a complex, polygenetic disorder, investigating brain morphometric correlates of PRS for AUD can refine the prediction of individuals at risk for problem drinking.

Sex differences have been extensively investigated in the clinical manifestation as well as the neurobiology and pathophysiology of AUD (Ceylan-Isik et al., 2010).

However, research aiming to identify sex differences in neural markers associated with genetic risks for alcohol dependence remains limited. Men relative to women have greater likelihood of early drinking, subsequent misuse, and higher lifetime prevalence of AUD (Flores-Bonilla and Richardson, 2020). Imaging studies appeared to show mixed results regarding the effects of sex on neurochemical, structural, and functional mechanisms associated with AUD (Verplaetse et al., 2021). A recent meta-analysis study revealed a group-by-sex interaction effect, with cerebellar GMVs more affected by AUD in women than in men and temporo-occipital and midcingulate GMVs more impacted in men as compared to women (Maggioni et al., 2023). An earlier study showed that male adolescents with vs. without family history of AUD had larger left hippocampal GMV, an effect not observed for females (Hanson et al., 2010). Left nucleus accumbens GMV was positively correlated with family history of AUD in girls but not in boys (Cservenka et al., 2015). These findings indicate potential sex differences in genetic risks for alcohol misuse. Addressing sex differences in the neural markers of PRS is crucial to our understanding of the sex-specific vulnerability to the development of AUD.

In the current study, we aimed at identifying volumetric correlates of alcohol use severity and PRS for alcohol dependence and examining the sex differences in a cohort of young adults studied in the Human Connectome Project (HCP). We evaluated drinking severity, computed PRS for each subject, and correlated GMVs of 166 regions in the automated anatomical atlas 3 (AAL3) with drinking severity and PRS, with age, sex, race, and total intracranial volume (TIV) as covariates in the same model. We also performed the analyses separately for men and women and, in post-hoc comparisons, employed slope tests to confirm sex differences. We broadly hypothesized shared and unique volumetric correlates of alcohol use severity and PRS for alcohol dependence as well as significant sex differences in these correlates.

## 2. Methods

### 2.1 Dataset: subjects and assessments

In the S1200 release, the HCP contains clinical, behavioral, and 3T magnetic resonance (MR) imaging data of 1,206 young adults (1,113 with structural MR scans) without severe neurodevelopmental, neuropsychiatric, or neurologic disorders. The current sample included 453 families with 1,140 healthy twins or siblings (ages 22-35; 618 women) who had data for genotyping. A total of 145 subjects were excluded from further analyses due to missing drinking metrics (n = 65) or imaging data (n = 80). The final sample size was 995 (534 women).

Participants were assessed with the Semi-Structured Assessment for the Genetics of Alcoholism (SSAGA), an instrument designed to assess physical, psychological and social manifestations of alcoholism and related disorders (Bucholz et al., 1994). As in our previous study (Li et al., 2022), we performed a principal component analysis on the 15 interrelated drinking metrics of the SSAGA in the HCP data, and identified one principal component (PC1) with an eigenvalue > 1 that accounted for 49.74% of the variance. The PC1 value reflected drinking severity (“PC1” henceforth) and served as the phenotype in the PRS analysis.

### 2.2 MRI protocol and voxel-based morphometry

A customized 3T Siemens Connectome Skyra with a standard 32-channel Siemens receiver head coil and a body transmission coil was used in the MRI scanning. T1-weighted high-resolution structural images were acquired using a 3D MPRAGE sequence with 0.7 mm isotropic resolution (FOV = 224 × 224 mm, matrix = 320 × 320, 256 sagittal slices, TR = 2,400 ms, TE = 2.14 ms, TI = 1,000 ms, FA = 8°).

As described in our prior work (Chen et al., 2022a, Chen and Li, 2022), we used voxel-based morphometry (VBM) to estimate the GMVs of brain regions with the CAT12 (Version 12.7) toolbox, following the suggested defaults of the CAT12 manual (Gaser and Dahnke, 2016): 1) individuals’ structural images were spatially normalized to the same stereotactic space; 2) the normalized images were segmented into gray matter, white matter, and cerebrospinal fluid; and 3) the gray matter (GM) images were smoothed. Specifically, a spatial adaptive non-local means (SANLM) denoising filter was first applied to the initial voxel-based processing (Manjón et al., 2010), followed by internal resampling to accommodate low-resolution images and anisotropic spatial resolutions. Subsequently, data were processed by bias-correction and affine-registration, followed by the unified segmentation (Ashburner and Friston, 2005). After segmentation, the brain was parcellated into left and right hemisphere, subcortical areas, and cerebellum after skull-stripping. A local intensity transformation of all tissue classes was performed to reduce the effects of higher GM intensities in the motor cortex, basal ganglia, or occipital lobe, followed by adaptive maximum a posteriori segmentation with partial volume estimation (Tohka et al., 2004). The tissue segments were then spatially normalized to a common reference space using DARTEL registrations (Ashburner, 2007). The GM maps were smoothed by convolution with an isotropic Gaussian kernel (FWHM = 8 mm). Data quality was checked by using the modules of display slices and VBM data homogeneity in the CAT12. The TIV estimated for each subject was used as a covariate in the following analyses.

### 2.3 Genotyping and PRS

The procedures of genotyping and PRS computation were described in detail in our prior work (Fu et al., 2024). Briefly, the discovery (base) sample included 14,904 unrelated cases with AUD (DSM-IV) and 37,944 unrelated healthy controls from the meta-analysis of ancestry-stratified genome-wide association (GWAS) of European and African-American cohorts by the Psychiatric Genomics Consortium (PGC) (Walters et al., 2018). The discovery sample was genotyped on microarrays and then imputed for 10,901,146 single nucleotide polymorphisms (SNPs). The current, HCP target sample was genotyped using Illumina Infinium Multi-Ethnic Genotyping Array (MEGA) or Infinium Neuro Consortium Array (2,292,654 SNPs). The computation of PRS requires base (i.e., discovery), target, phenotype, and covariate data (Choi et al., 2020). The summary statistics of GWAS served as the base data, and the whole-genome genotype and phenotype data (i.e., PC1) of the HCP sample served as the target data.

#### 2.3.1 Quality control for both base and target samples

We performed quality control on both base and target data based on a standard protocol (Zuo et al., 2012, Choi et al., 2020), to filter out the SNPs with ambiguous alleles or those mismatched between base and target data, and to exclude the subjects with sex mismatching between self-report and sex chromosomes. We also removed the samples with genetic relatedness for any pair of individuals between base and target samples. Finally, we checked the QQ plot of all *p* values from the cleaned base dataset and estimated the genomic inflation factor (λ), to further filter out the SNP outliers until the genomic inflation remained reasonably low. Only the SNPs present in both “cleaned” base and target samples were included for PRS computation. The final base data had a *λ* = 1.04 and chip-heritability > 0.05, and the risk alleles all had a *Z*-score > 1 for the effect size of SNP-AUD associations.

#### 2.3.2 PRS calculation

PRS was computed according to a formula introduced by Choi et al. (Choi et al., 2020) that considered the effect size of SNPs, the number of risk alleles, and the total number of SNPs included (n = 4 with *p* threshold < 5e-08). In brief, PRS was computed as a sum of the genome-wide risk alleles, weighted by the corresponding effect size estimated from GWAS. We calculated the PRS for each subject using the program PRSice-2 (Choi et al., 2020), which automatically excluded the correlated SNPs (pairwise *r*^2^ > 0.25) and considered age, sex (for all), and principal components of ancestry as covariates. The best-fit PRS with the best discriminative capacity was determined based on the highest area under the receiver-operator curve (AUC) in a logistic regression model with the phenotype as the outcome and the PRS, age, sex (when computed for males and females together), and ancestry proportions as covariates (Khera et al., 2018).

To ensure that the subjects were unrelated within each target data subset, as required for the computation of the PRS, the twins and siblings of each family in HCP samples were randomly separated into independent target subsets. To maximize the sample size of each subset, the subjects within the first subset who were unrelated to the subjects of the other subsets were filled in each of the latter subsets repeatedly. The PRS and the percentage of phenotypic variation explained by the PRS (i.e., *R*^2^) were then calculated separately for each subset under the same p-value thresholds. The PRS of the same subject, if showing a difference < 1% across the subsets, were averaged as the final PRS. The percentages of phenotypic variation, if showing a difference < 1% across the subsets, were averaged as the final percentages.

With age, sex (for all), and race as covariates, we computed the correlation coefficients between PC1 and PRS across all subjects as well as in men and women separately.

### 2.4 Region of interest (ROI) analyses of GMV

We estimated the GMVs of 166 regions in the AAL3 (**Supplementary Table S1**) (Rolls et al., 2020) and computed the correlation coefficients of regional GMVs with PC1 and PRS in a single regression model, controlling for age, sex (for all), race, and TIV in men and women combined and separately. For the correlations that were significant for either men or women alone, we also performed slope tests to examine sex differences in the correlations (Chen et al., 2021, Li et al., 2020). Note that this analysis did not represent “double-dipping” as a correlate identified in men may have just missed the threshold in women and vice versa. Thus, slope tests were needed to confirm sex differences.

In addition to the ROI analyses, we also performed whole-brain regressions against PC1 and PRS in a single model to identify the volumetric correlates, with age, sex (for all), race, and TIV as covariates, in all and in men and women alone. We evaluated the results at voxel *p* < 0.001, uncorrected along with cluster *p* < 0.05 corrected for family-wise error (FWE) of multiple comparisons, based on the Gaussian random field theory, as implemented in the SPM12. However, we did not observe any significant clusters showing correlations with PC1 or with PRS in men and women combined or separately in the whole-brain analyses. Thus, we presented the results of ROI analyses on the 166 regions of AAL3 in the below.

## 3. Results

### 3.1 Demographic and clinical measures

The descriptive statistics and *t/χ²* tests of sex differences in age, race, PC1, PRS, and TIV are shown in **Table 1**. Women were older than men (*t* = 7.32, *p* < 0.001). There was no significant sex difference in race composition (*χ*² = 3.22, *p* = 0.666). With age and race as covariates, men vs. women showed significantly higher PC1, PRS, and TIV (*t’s* ≥ 11.07, *p’s* ≤ 0.015).

**Table 1.** Demographic and clinical measures of men and women.

| | Men (n = 461) | Women (n = 534) | $t/\chi^2$ | $p$ |
| --- | --- | --- | --- | --- |
| Age (years) | 27.86 ± 3.58 | 29.53 ± 3.61 | -7.32 | < 0.001 |
| Race |  |  | 3.22 | 0.666 |
| AI/AN | 1 (0.22%) | 4 (0.75%) |  |  |
| A/NH/OPI | 25 (5.42%) | 28 (5.24%) |  |  |
| AA | 55 (11.93%) | 84 (15.73%) |  |  |
| White | 360 (78.09%) | 396 (74.16%) |  |  |
| > one race | 12 (2.60%) | 15 (2.81%) |  |  |
| UK/NR | 8 (1.74%) | 11 (2.06%) |  |  |
| Drinking PC1 | 0.39 ± 1.07 | -0.34 ± 0.79 | 11.01 | < 0.001 |
| PRS | 3.49 ± 0.80 | 3.38 ± 0.82 | 2.44 | 0.015 |
| TIV (ccm) | 1620.7 ± 157.2 | 1392.2 ± 125.0 | 23.03 | < 0.001 |

*Note:* Values are mean ± SD or n (%). AI/AN: American Indian/Alaskan Native; A/NH/OPI: Asian/Native Hawaiian/Other Pacific Islander; AA: African American; UK/NR: Unknown/Not Reported; PC1: first principal component of drinking metrics; PRS: polygenic risk score. A $\chi^2$ test was used to examine sex differences in race. Independent $t$ tests for PC1, PRS, and TIV were performed with age and race as covariates.

With age, sex (for all), and race as covariates, PC1 and PRS were significantly correlated in all subjects (*r* = 0.152, *p* = 0.000002) as well as in men (*r* = 0.130, *p* = 0.005235) and women (*r* = 0.174, *p* = 0.000055) separately. The slope test showed that the sex difference in the correlations was not significant (*t* = 0.114, *p* = 0.908915).

### 3.2 Volumetric correlates of PC1 and PRS

Figure 1 shows the volumetric correlates of PC1 and PRS in all, men, and women, with PC1 and PRS modeled together and age, sex (for all), race, and TIV as covariates. Specifically, for all subjects, higher PC1 was correlated with lower GMVs of the right insula (INS; *r* =-0.126, *p* = 0.000068) and Heschl’s gyrus (HES; *r* =-0.126, *p* = 0.000074). Higher PRS was correlated with higher GMVs of left posterior orbitofrontal gyrus (OFGp; *r* = 0.128, *p* = 0.000053), and bilateral intralaminar nuclei of the thalamus (IL-THA; left: *r* = 0.147, *p* = 0.000003 and right: *r* = 0.144, *p* = 0.000005) and lingual gyri (LG; left: *r* = 0.132, *p* = 0.000030 and right: *r* = 0.125, *p* = 0.000077). No regions showed positive correlations with PC1 or negative correlation with PRS at the same threshold. **Table 2** summarizes these volumetric correlates for both PC1 and PRS as well as the slope test results. With a *p*-value of 0.007 (0.05/7) to correct for multiple comparisons, all volumetric correlates were unique to either PC1 or PRS, except for the left lingual gyrus.

**Figure 1.**
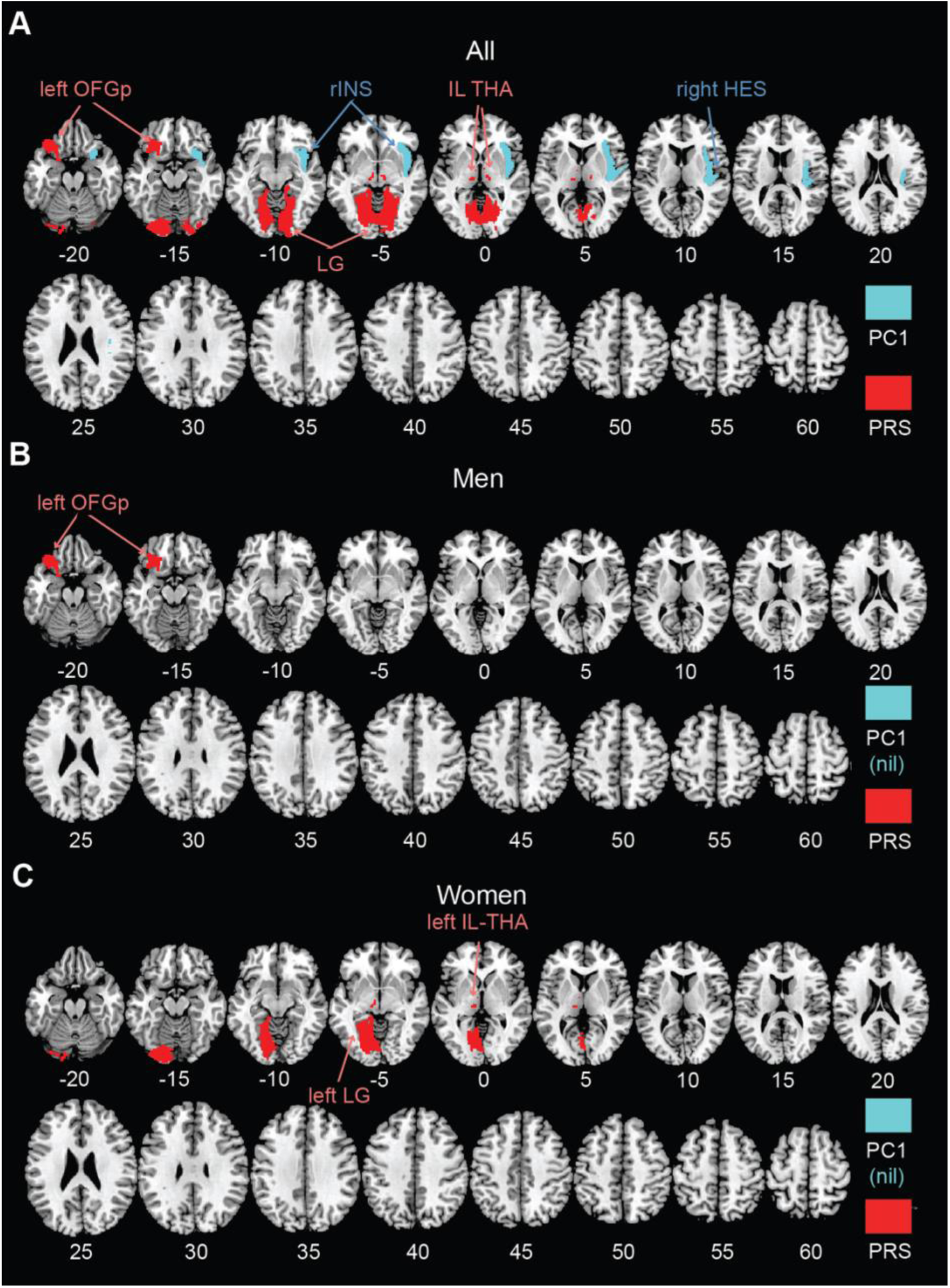
AAL3 regional GMVs in significant correlation with PC1 (cyan; negative) and PRS (red; positive) in a single regression model for **(A)** all, **(B)** men, and **(C)** women, with age, sex (for all), race, and TIV as covariates. No regions showed positive correlations with PC1 or negative correlation with PRS at the same threshold. PC1: first principal component of drinking metrics; PRS: polygenic risk score; OFGp: posterior orbitofrontal gyrus; INS: insula; IL-THA: intralaminar nucleus of thalamus; HES: Heschl’s gyrus; LG: lingual gyrus; nil: no significant findings.

**Table 2.** Correlations of volumetric correlates with PC1 and PRS and slope tests.

| GMV | PC1<br>*r* | PRS<br>*r* | Slope test<br>*t* | *p* |
| --- | --- | --- | --- | --- |
| **All** |  |  |  |  |
| right INS | **-0.126** | 0.027 | 3.599 | 3.2733e-04* |
| right HES | **-0.126** | 0.007 | 3.181 | 0.0015* |
| left OFGp | -0.016 | **0.128** | 2.985 | 0.0029* |
| left IL-THA | -0.094 | **0.147** | 5.276 | 1.4612e-07* |
| right IL-THA | -0.056 | **0.144** | 4.289 | 1.8772e-05* |
| left LG | 0.015 | **0.132** | 2.325 | 0.0202 |
| right LG | -0.019 | **0.125** | 2.992 | 0.0028* |
| **Men** |  |  |  |  |
| left OFGp | -0.011 | **0.200** | 2.691 | 0.0073 |
| **Women** |  |  |  |  |
| left IL-THA | -0.033 | **0.185** | 3.577 | 3.6295e-04* |
| left LG | -0.005 | **0.166** | 2.816 | 0.0050* |

*Note:* GMV: gray matter volumes; PC1: first principal component of drinking metrics; PRS: polygenic risk score; OFGp: posterior orbitofrontal gyrus; INS: insula; IL-THA: intralaminar nucleus of thalamus; HES: Heschl's gyrus; LG: lingual gyrus. Bold $r$ values represent significant correlations at corrected $p < 0.05/(166 \times 2) = 0.00015$ . \* $p < 0.05/7 = 0.00714$ for multiple corrections on slope tests.

For men or women considered separately, PC1 was not significantly correlated with the GMVs of any AAL3 regions. In men, higher PRS was associated with higher GMV in the left OFGp (*r* = 0.200, *p* = 0.000016). In women, higher PRS was associated with higher GMVs in the left IL-THA (*r* = 0.185, *p* = 0.000019) and LG (*r* = 0.166, *p* = 0.000120). Moreover, slope tests confirmed that, in women, the correlations of PRS with left IL-THA GMV (*t* = 3.577, *p* = 3.6295e-04) and with left LG GMV (*t* = 2.816, *p* = 0.0050) were stronger than those with PC1. The correlation of the left OFGp GMV with PRS was also stronger than that with PC1 in men, although the p value (0.0073) just missed the corrected threshold. However, slope tests did not show significant sex differences in any of these GMV correlations with PRS (*t*‘s ≤ 1.421, *p*‘s ≥ 0.0729).

## 4. Discussion

We aimed to distinguish regional volumetric markers of the genetic risks, as indexed by the PRS, of alcohol misuse from those reflecting the effects of alcohol exposure, as indexed by drinking severity PC1. To this end we employed a single regression model where both PRS and PC1 served as independent variables or predictors, so that the effects of genetic risks and alcohol exposure were mutually accounted for. We observed distinct volumetric correlates of PRS and PC1 and discussed the main findings in the below.

### 4.1 Volumetric correlates of drinking severity

We identified gray matter atrophy in the right insula and Heschl’s gyrus in relation to drinking severity, in line with prior reports (Taki et al., 2006). Insular GMV loss has been observed in drinkers (Grodin et al., 2017), smokers (Chen et al., 2022a), and other drug users (Naqvi et al., 2014) who often engage in heavy alcohol use. A critical hub of the interoceptive system, the insula is affected by alcohol’s neurotoxicity both structurally and functionally, and insula dysfunction contributes to the development of addiction (Wisniewski et al., 2021). The current finding of insular GMV loss in a sample of largely non-dependent drinkers suggested that volumetric reduction of the insula can transpire early in the course of alcohol use.

Notably, a recent study with the HCP data demonstrated significant negative correlations between alcohol consumption and GMVs of right insula and middle/superior frontal gyri and attributed these volumetric alterations to shared genetic factors, based on the correlations of monozygotic vs. dizygotic twins (Baranger et al., 2020). In the latter study, a concordant sibling pair was defined as a pair who were both in the same category of consumption (i.e., high or low). Conversely, a pair was considered discordant if the siblings belonged to different consumption categories. Siblings in the concordant high and low group each had lowest and highest insula and frontal GMVs, respectively. On the other hand, the GMVs did not differ between low and high alcohol-using siblings within the discordant pairs. The findings suggested genetic risks rather than alcohol use as the primary driver of GMV reductions. In contrast to our study, however, the latter study did not directly control for the influence of alcohol use when investigating the relationship between genetic risks and insula GMV. Alcohol exposure may play a role in shaping the association between insula GMV and genetic risks. Insular GMV as a genetic risk marker of alcohol misuse needs to be revisited in future studies, including those of patients with AUD.

The Heschl’s gyrus supports auditory and semantic processing (Schneider et al., 2002, Khalighinejad et al., 2021) but its role in substance misuse is less clear.

Consistent with our findings, lower GMV in the left Heschl’s gyrus was associated with higher drinking severity in young and middle-aged adults (Thayer et al., 2016). A few meta-analyses also showed GMV deficits in Heschl’s gyrus in patients with other substance misuse, including opioid (Wollman et al., 2017) and cocaine (Hall et al., 2015) dependence and misuse of multiple substances (Zhang et al., 2021), where heavy drinking has frequently been observed.

### 4.2 Volumetric correlates of the genetic risks of AUD

We computed PRS using the GWAS data for AUD and found that the PRS was significantly associated with drinking severity in our sample of young adults. With drinking severity accounted for, we found PRS in positive correlation with regional GMV’s in left posterior orbitofrontal gyrus, bilateral intralaminar nuclei of the thalamus, and lingual gyri. These volumetric correlates may represent a genetically informed risk marker of alcohol misuse. A recent Adolescent Brain Cognition Development study showed that higher PRS for problem alcohol use was associated with greater cortical thickness of the right supramarginal gyrus in substance-naïve children (Hatoum et al., 2021). Substance-naïve offsprings of AUD patients have been found to have thicker frontal and temporo-parietal cortices (Holla et al., 2019), and many of these cortical regions are thinner in individuals with alcohol dependence (Mackey et al., 2019b).

Although GMV and cortical thickness are different neural metrics, these previous reports are consistent with the current findings of distinct volumetric markers of PRS and severity of alcohol misuse.

It has been postulated that greater cortical thickness among those at familial risk for alcohol problems reflects a developmental delay in neural pruning, placing children and adolescents at risk for substance and alcohol use, which, in turn, accelerates neuronal pruning in later development. While no other studies to our knowledge have specifically reported larger regional volumes in link with the generic vulnerability of alcohol misuse, investigators have observed similar findings in clinical conditions comorbid with AUD. For instance, greater PRS for depression was associated with larger GMVs in bilateral hippocampi and right gyrus rectus (Fu et al., 2024) and right inferior and middle temporal gyri (Wang et al., 2023). It remains to be seen whether the comorbid conditions may share genetic bases.

Structural changes of the thalamus have long been documented in problem drinkers (Ide et al., 2017, Yang et al., 2016, Mon et al., 2014). In a study of meta-analysis, patients with AUD demonstrated significant GMV reduction in the left thalamus as compared to non-drinking controls, with the severity of reduction correlated with life-time alcohol consumption (Yang et al., 2016). No prior research has addressed whether thalamic subnuclei may be differentially impacted by alcohol or genetic risks of alcohol misuse. Here, we observed that bilateral intralaminar nuclei of the thalamus (IL-THA) GMVs were positively correlated with PRS, in contrast to volumetric reduction as observed for the thalamus as a result of chronic alcohol use. The IL-THA projects to cortical regions, dorsal and ventral striatum, and the amygdala, and supports physiological arousal, vigilance, and motivated behavior (Vertes et al., 2015). If replicated, these findings suggest that specific thalamic subnuclei, such as the IL-THA, may play a critical role in the neurobiological mechanisms underlying genetic risks for alcohol misuse. Studies combining genetic analyses and brain imaging can assess how intralaminar thalamic dysfunction may support ill-directed arousal to reward and/or negative emotions, disposing individuals to earlier and heavier drinking.

The orbitofrontal gyrus (OFG) supports goal-directed behaviors particularly through evaluation of rewards and other outcomes, which have been shown to be disrupted in substance misuse, including AUD (Moorman, 2018). Within the OFG, the posterior region is primarily involved in emotion perception and processing (Barbas, 2007, Timbie and Barbas, 2015). Chronic alcohol use was associated with reduced OFG volumes (McCalley and Hanlon, 2021). In addition, OFG function appears to be influenced by genetic variations in neurotransmitter receptors, such as the A-G allele variants in the OPRM1 receptor and the 7-repeat allele of the DRD4 receptor. For example, the OFG shows higher response to alcohol relative to neutral cues in heavy drinkers, with stronger activation observed in those carrying the OPRM1G allele, suggesting a genetic predispositions to alcohol dependence (Filbey et al., 2008, Ray and Hutchison, 2004). Other studies reported greater neuroinflammation in the OFG (Vetreno et al., 2013), which may contribute to changes in the OFG volumes (Jones et al., 2019, Wang et al., 2023), in individuals with AUD. Although it is premature to attempt to reconcile these findings, the studies together suggest the importance of distinguishing genetic risks in the investigation of the pathophysiology of AUD.

The lingual gyrus, located in the occipital lobe and involved in visual processing (Spagna et al., 2023), has been implicated in alcohol misuse in prior research. For instance, heightened activation in the bilateral lingual gyri has been associated with alcohol cue-induced craving among individuals with AUD (Park et al., 2007).

Interestingly, enlargement of visual processing regions, including the lingual gyrus, has been observed in anxiety and depressive disorders, which are common comorbidities of AUD (Frick et al., 2014, Jung et al., 2014). Additionally, we recently demonstrated stronger resting-state functional connectivity between the subgenual anterior cingulate cortex and the lingual gyrus in association with higher PRS for depression (Chen et al., 2025). Future research should explore both structural and functional characteristics of the lingual gyrus in relation to genetic predisposition for alcohol dependence.

### 4.3 Potential sex differences?

We did not observe significant sex difference in PRS-PC1 correlation or in the volumetric correlates of PC1 or PRS, despite the finding of both higher PRS and PC1 in men vs. women. Earlier family, adoption, and twin studies found similar heritability estimates of alcohol dependence (Tawa et al., 2016) and GMVs (Chen et al., 2022b) across sexes. However, the patterns of morphological brain deficits as a result of chronic drinking appeared to differ between men and women (Pfefferbaum et al., 2001, Squeglia et al., 2012, Maggioni et al., 2023, Rossetti et al., 2021, Grace et al., 2021).

Moreover, in alcohol-naïve adolescents with a family history of AUD, males exhibited larger left hippocampal GMV, a pattern that was not observed in their female counterparts (Hanson et al., 2010). In another study, healthy female but not male adolescents with a family history of AUD showed larger GMV of the left nucleus accumbens (Cservenka et al., 2015). These findings suggest potential sex differences in the brain structure related to the genetic predisposition to alcohol misuse, as has been demonstrated for sex differences in brain functioning, e.g., the roles of dopaminergic signaling in risk/novelty seeking (Berman et al., 2003) and hormonal modulation of dopaminergic signaling (Ceylan-Isik et al., 2010), that may dispose individuals to early and more severe drinking. Further, GMV correlates of clinical conditions comorbid with problem drinking, including ADHD (Luo et al., 2023, Luo et al., 2022a) and depression (Fu et al., 2024) or of psychological constructs that conduce to problem drinking, including impulsivity (Chen et al., 2022b, Ide et al., 2020), manifest robust sex differences. Studies of larger sample sizes and with individuals with more severe drinking problems are needed to better understand how sex differences are reflected in the risk markers or inter-related outcomes of chronic alcohol consumption.

### 4.4 Limitations and conclusions

There are several limitations to consider. First, HCP participants were healthy (“neurotypical”) young adults without severe drinking problems; drinkers that required treatment were excluded in recruitment. Thus, the current findings should be regarded as specific to this sample. In particular, no ROIs showed GMVs in negative correlation with PC1 in men or in women alone, a finding that likely reflects this young adult, largely social-drinking population. Second, we did not consider other substance uses in our analyses. Nicotine or other drug use may have additive or interactive effects on regional GMVs (Grodin et al., 2021, Luo et al., 2022b). Third, longitudinal studies are needed to better understand the distinct impacts of genetic risks and alcohol on the brain.

In conclusion, we replicated GMV reduction in association with greater severity of alcohol use and provided evidence for regional GMVs in positive association with the genetic risks of alcohol dependence in young healthy adults. Our findings suggest potentially unique volumetric markers for severity of drinking and genetic risks for alcohol dependence.

## CRediT authorship contribution statement

**Yu Chen:** Conceptualization, Methodology, Software, Data Curation, Formal Analysis, Writing – Original Draft, Writing – Review & Editing, Visualization. **Huey-Ting Li:** Software, Data Curation, Formal Analysis, Writing – Original Draft, Writing – Review & Editing. **Xingguang Luo:** Methodology, Software, Data Curation, Formal Analysis, Writing – Original Draft, Writing – Review & Editing. **Guangfei Li:** Formal Analysis, Writing – Original Draft, Writing – Review & Editing. **Jaime S. Ide:** Formal Analysis, Writing – Original Draft, Writing – Review & Editing. **Chiang-Shan R. Li:** Conceptualization, Methodology, Writing – Original Draft, Writing – Review & Editing, Supervision, Funding Acquisition.

## Data Availability

https://www.humanconnectome.org/study/hcp-young-adult/data-releases

## Acknowledgements

This study is supported by NIH grant AG072893. The NIH is otherwise not responsible for the conceptualization of the study, data collection and analysis, or in the decision in publishing the results.

## Data availability, Ethics declarations, and Consent to participate

We have obtained permission from the Human Connectome Project (HCP) to use the Open and Restricted Access data for the current study. Data were provided by the WU-Minn Consortium (Principal Investigators: David Van Essen and Kamil Ugurbil; 1U54MH091657) funded by the 16 NIH Institutes and Centers that support the NIH Blueprint for Neuroscience Research; and by the McDonnell Center for Systems Neuroscience at Washington University. The HCP young-adult data is publicly available at https://www.humanconnectome.org/study/hcp-young-adult/.

