## Supplementary Materials for "The effects of alcohol use severity and polygenic risk on gray matter volumes in young adults"

**Supplementary Table S1.** Abbreviations and labels of parcellations in the AAL3 atlas

| # | Abbreviation | Label |
|---|---|---|
| 1 | lPreCG | Left Precentral gyrus |
| 2 | rPreCG | Right Precentral gyrus |
| 3 | lSFG | Left Superior frontal gyrus-dorsolateral |
| 4 | rSFG | Right Superior frontal gyrus-dorsolateral |
| 5 | lMFG | Left Middle frontal gyrus |
| 6 | rMFG | Right Middle frontal gyrus |
| 7 | lIFGoperc | Left Inferior frontal gyrus-opercular part |
| 8 | rIFGoperc | Right Inferior frontal gyrus-opercular part |
| 9 | lIFGtriang | Left Inferior frontal gyrus-triangular part |
| 10 | rIFGtriang | Right Inferior frontal gyrus-triangular part |
| 11 | lIFGorb | Left IFG pars orbitalis |
| 12 | rIFGorb | Right IFG pars orbitalis |
| 13 | lROL | Left Rolandic operculum |
| 14 | rROL | Right Rolandic operculum |
| 15 | lSMA | Left Supplementary motor area |
| 16 | rSMA | Right Supplementary motor area |
| 17 | lOLF | Left Olfactory cortex |
| 18 | rOLF | Right Olfactory cortex |
| 19 | lSFGmedial | Left Superior frontal gyrus-medial |
| 20 | rSFGmedial | Right Superior frontal gyrus-medial |
| 21 | lPFCventmed | Left Superior frontal gyrus-medial orbital |
| 22 | rPFCventmed | Right Superior frontal gyrus-medial orbital |
| 23 | lREC | Left Gyrus rectus |
| 24 | rREC | Right Gyrus rectus |
| 25 | lOFCmed | Left Medial orbital gyrus |
| 26 | rOFCmed | Right Medial orbital gyrus |
| 27 | lOFCant | Left Anterior orbital gyrus |
| 28 | rOFCant | Right Anterior orbital gyrus |
| 29 | lOFCpost | Left Posterior orbital gyrus |
| 30 | rOFCpost | Right Posterior orbital gyrus |
| 31 | lOFClat | Left Lateral orbital gyrus |
| 32 | rOFClat | Right Lateral orbital gyrus |
| 33 | lINS | Left Insula |
| 34 | rINS | Right Insula |
| 37 | lMCC | Left Middle cingulate & paracingulate gyri |
| 38 | rMCC | Right Middle cingulate & paracingulate gyri |

| | | |
|---|---|---|
| 39 | lPCC | Left Posterior cingulate gyrus |
| 40 | rPCC | Right Posterior cingulate gyrus |
| 41 | lHIP | Left Hippocampus |
| 42 | rHIP | Right Hippocampus |
| 43 | lPHG | Left Parahippocampal gyrus |
| 44 | rPHG | Right Parahippocampal gyrus |
| 45 | lAMYG | Left Amygdala |
| 46 | rAMYG | Right Amygdala |
| 47 | lCAL | Left Calcarine fissure and surrounding cortex |
| 48 | rCAL | Right Calcarine fissure and surrounding cortex |
| 49 | lCUN | Left Cuneus |
| 50 | rCUN | Right Cuneus |
| 51 | lLING | Left Lingual gyrus |
| 52 | rLING | Right Lingual gyrus |
| 53 | lSOG | Left Superior occipital gyrus |
| 54 | rSOG | Right Superior occipital gyrus |
| 55 | lMOG | Left Middle occipital gyrus |
| 56 | rMOG | Right Middle occipital gyrus |
| 57 | lIOG | Left Inferior occipital gyrus |
| 58 | rIOG | Right Inferior occipital gyrus |
| 59 | lFFG | Left Fusiform gyrus |
| 60 | rFFG | Right Fusiform gyrus |
| 61 | lPoCG | Left Postcentral gyrus |
| 62 | rPoCG | Right Postcentral gyrus |
| 63 | lSPG | Left Superior parietal gyrus |
| 64 | rSPG | Right Superior parietal gyrus |
| 65 | lIPG | Left Inferior parietal gyrus-excluding supramarginal and angular gyri |
| 66 | rIPG | Right Inferior parietal gyrus-excluding supramarginal and angular gyri |
| 67 | lSMG | Left SupraMarginal gyrus |
| 68 | rSMG | Right SupraMarginal gyrus |
| 69 | lANG | Left Angular gyrus |
| 70 | rANG | Right Angular gyrus |
| 71 | lPCUN | Left Precuneus |
| 72 | rPCUN | Right Precuneus |
| 73 | lPCL | Left Paracentral lobule |
| 74 | rPCL | Right Paracentral lobule |
| 75 | lCAU | Left Caudate nucleus |
| 76 | rCAU | Right Caudate nucleus |
| 77 | lPUT | Left Lenticular nucleus-Putamen |
| 78 | rPUT | Right Lenticular nucleus-Putamen |
| 79 | lPAL | Left Lenticular nucleus-Pallidum |
| 80 | rPAL | Right Lenticular nucleus-Pallidum |
| 83 | lHES | Left Heschls gyrus |

| | | |
|---|---|---|
| 84 | rHES | Right Heschls gyrus |
| 85 | lSTG | Left Superior temporal gyrus |
| 86 | rSTG | Right Superior temporal gyrus |
| 87 | lTPOsup | Left Temporal pole: superior temporal gyrus |
| 88 | rTPOsup | Right Temporal pole: superior temporal gyrus |
| 89 | lMTG | Left Middle temporal gyrus |
| 90 | rMTG | Right Middle temporal gyrus |
| 91 | lTPOmid | Left Temporal pole: middle temporal gyrus |
| 92 | rTPOmid | Right Temporal pole: middle temporal gyrus |
| 93 | lITG | Left Inferior temporal gyrus |
| 94 | rITG | Right Inferior temporal gyrus |
| 95 | lCERCRU1 | Left Crus I of cerebellar hemisphere |
| 96 | rCERCRU1 | Right Crus I of cerebellar hemisphere |
| 97 | lCERCRU2 | Left Crus II of cerebellar hemisphere |
| 98 | rCERCRU2 | Right Crus II of cerebellar hemisphere |
| 99 | lCER3 | Left Lobule III of cerebellar hemisphere |
| 100 | rCER3 | Right Lobule III of cerebellar hemisphere |
| 101 | lCER4_5 | Left Lobule IV-V of cerebellar hemisphere |
| 102 | rCER4_5 | Right Lobule IV-V of cerebellar hemisphere |
| 103 | lCER6 | Left Lobule VI of cerebellar hemisphere |
| 104 | rCER6 | Right Lobule VI of cerebellar hemisphere |
| 105 | lCER7b | Left Lobule VIIB of cerebellar hemisphere |
| 106 | rCER7b | Right Lobule VIIB of cerebellar hemisphere |
| 107 | lCER8 | Left Lobule VIII of cerebellar hemisphere |
| 108 | rCER8 | Right Lobule VIII of cerebellar hemisphere |
| 109 | lCER9 | Left Lobule IX of cerebellar hemisphere |
| 110 | rCER9 | Right Lobule IX of cerebellar hemisphere |
| 111 | lCER10 | Left Lobule X of cerebellar hemisphere |
| 112 | rCER10 | Right Lobule X of cerebellar hemisphere |
| 113 | ltAV | Left Thalamus-Anteroventral Nucleus |
| 114 | rtAV | Right Thalamus-Anteroventral Nucleus |
| 115 | ltLP | Left Lateral posterior |
| 116 | rtLP | Right Lateral posterior |
| 117 | ltVA | Left Ventral anterior |
| 118 | rtVA | Right Ventral anterior |
| 119 | ltVL | Left Ventral lateral |
| 120 | rtVL | Right Ventral lateral |
| 121 | ltVPL | Left Ventral posterolateral |
| 122 | rtVPL | Right Ventral posterolateral |
| 123 | ltIL | Left Intralaminar |
| 124 | rtIL | Right Intralaminar |
| 125 | ltRe | Left Reuniens |
| 126 | rtRe | Right Reuniens |

| | | |
|---|---|---|
| 127 | ltMDm | Left Mediodorsal medial magnocellular |
| 128 | rtMDm | Right Mediodorsal medial magnocellular |
| 129 | ltMDl | Left Mediodorsal lateral parvocellular |
| 130 | rtMDl | Right Mediodorsal lateral parvocellular |
| 131 | ltLGN | Left Lateral geniculate |
| 132 | rtLGN | Right Lateral geniculate |
| 133 | ltMGN | Left Medial Geniculate |
| 134 | rtMGN | Right Medial Geniculate |
| 135 | ltPuA | Left Pulvinar anterior |
| 136 | rtPuA | Right Pulvinar anterior |
| 137 | ltPuM | Left Pulvinar medial |
| 138 | rtPuM | Right Pulvinar medial |
| 139 | ltPuL | Left Pulvinar lateral |
| 140 | rtPuL | Right Pulvinar lateral |
| 141 | ltPuI | Left Pulvinar inferior |
| 142 | rtPuI | Right Pulvinar inferior |
| 143 | lACCsub | Left Anterior cingulate cortex-subgenual |
| 144 | rACCsub | Right Anterior cingulate cortex-subgenual |
| 145 | lACCpre | Left Anterior cingulate cortex-pregenual |
| 146 | rACCpre | Right Anterior cingulate cortex-pregenual |
| 147 | lACCsup | Left Anterior cingulate cortex-supracallosal |
| 148 | rACCsup | Right Anterior cingulate cortex-supracallosal |
| 149 | lNacc | Left Nucleus accumbens |
| 150 | rNacc | Right Nucleus accumbens |
| 151 | lVTA | Left Ventral tegmental area |
| 152 | rVTA | Right Ventral tegmental area |
| 153 | lSNpc | Left Substantia nigra-pars compacta |
| 154 | rSNpc | Right Substantia nigra-pars compacta |
| 155 | lSNpr | Left Substantia nigra-pars reticulata |
| 156 | rSNpr | Right Substantia nigra-pars reticulata |
| 157 | lRedN | Left Red nucleus |
| 158 | rRedN | Right Red nucleus |
| 159 | lLC | Left Locus coeruleus |
| 160 | rLC | Right Locus coeruleus |
| 161 | VER1_2 | Lobule I-II of vermis |
| 162 | VER3 | Lobule III of vermis |
| 163 | VER4_5 | Lobule IV-V of vermis |
| 164 | VER6 | Lobule VI of vermis |
| 165 | VER7 | Lobule VII of vermis |
| 166 | VER8 | Lobule VIII of vermis |
| 167 | VER9 | Lobule IX of vermis |
| 168 | VER10 | Lobule X of vermis |
| 169 | RapheD | Raphe nucleus-dorsal |

| | | |
|---|---|---|
| 170 | RapheM | Raphe nucleus-median |

*Note:* The original numbering in AAL2 for the anterior cingulate cortex (35, 36) and thalamus (81, 82) is left empty in AAL3 because finer parcellations of these regions are provided in AAL3. The total number of parcellations in AAL3 is 166, with the maximum label number 170 (Rolls, Huang, Lin, Feng, & Joliot, 2020).
